# Immunological Peculiarities of Granulomatosis with Polyangiitis and Eosinophilic Granulomatosis with Polyangiitis in a Tunisian Cohort

**DOI:** 10.64898/2026.11.18.26368222

**Authors:** Sana Toujani, Youssef Belhaj Hassen, Amira El Ouni, Asma Belhassen, Cherifa Abdelkefi, Asma Tekaya, Thara Larbi, Zeineb Meddeb, Salwa Hamzaoui

## Abstract

**Background:** Granulomatosis with polyangiitis (GPA) and eosinophilic granulomatosis with polyangiitis (EGPA) are among the rarest systemic vasculitides. They are characterised by variable presentation, necrotizing and granulomatous vasculitis, and inconstant ANCA positivity. Objective. We aimed to study the clinical and immunological profile of GPA and EGPA in a Tunisian cohort.

**Methods:** A descriptive, retrospective, monocentric study over 22 years (2000–2021). GPA and EGPA cases were recorded in an internal-medicine department. Diagnosis was made according to the ACR 1990 criteria, and/or the 2012 Chapel Hill Consensus Conference, and/or the Diagnostic and Classification Criteria for Vasculitis Study (DCVAS 2018).

**Results:** We included 19 GPA and 11 EGPA patients, with respective median ages of 54 and 52 years and a female predominance. Late-onset asthma and hypereosinophilia were constant in EGPA. GPA had a predominant generalized form; we identified anti-MPO (58%), anti-PR3 (26.5%), dual antigenic specificity (11%) and atypical x-ANCA (5.5%). ANCA were negative in EGPA (55%) or directed against MPO. ANCA positivity was associated with peripheral neuropathy (p=0.013) in EGPA. MPO-ANCA were associated with female sex (p=0.005) and anti-PR3 with male sex (p=0.018), smoking (p=0.019), multiplicity of organs affected (p=0.006) and gastrointestinal involvement (p=0.033). Under conventional treatment, 36% of EGPA and 42% of GPA patients relapsed. Mortality was 11% in GPA, correlated with a higher BVAS version 3 at diagnosis (p=0.012), heart block (p=0.039) and neurological damage (p=0.050).

**Conclusion:** We provide a detailed profile of GPA and EGPA in a Tunisian cohort. The high prevalence of anti-MPO GPA and the presence of dual positivity suggest a North–South gradient in ANCA specificity and justify larger multicentre studies of the immunological, genetic and environmental determinants of this specificity.

## Background

Granulomatosis with polyangiitis (GPA) and eosinophilic granulomatosis with polyangiitis (EGPA), formerly Wegener granulomatosis and Churg–Strauss syndrome, are among the rarest systemic vasculitides, with a recent meta-analysis estimating a pooled incidence of anti-neutrophil cytoplasmic antibody (ANCA)-associated vasculitis (AAV) of 17.2 per million person-years and a prevalence of 198 per million [1]. Their positive diagnosis rests on a combination of clinical features, histological evidence of necrotizing and granulomatous vasculitis, and the inconstant positivity of ANCA [2].

ANCA are detected in the large majority of patients with GPA, whereas approximately 60% of EGPA cases are ANCA-negative [3]. ANCA are classically directed against proteinase 3 (PR3) in GPA and against myeloperoxidase (MPO) in microscopic polyangiitis (MPA) and EGPA, but there is a great variability in the clinical and immunological profile of both diseases [4, 5].

Importantly, genetic and clinical data increasingly indicate that ANCA antigen specificity, rather than the clinical syndrome alone, captures the underlying biology of AAV. PR3-ANCA and MPO-ANCA disease may therefore represent distinct autoimmune entities, with EGPA further divisible by ANCA status [6–8]. Against this background, we analysed the clinical characteristics of GPA and EGPA according to their immunological profile in a Tunisian cohort.

## Methods

We conducted a descriptive, retrospective, monocentric study over a period of twenty-two years. We collected all cases of GPA and EGPA diagnosed between 2000 and 2019 in an internal-medicine department. Patients were followed until December 2021. The diagnosis was made with reference to the American College of Rheumatology criteria (ACR 1990) [9, 10], and/or the Chapel Hill Consensus Conference (CHCC 2012) [2], and/or the Diagnostic and Classification Criteria for Vasculitis Study (DCVAS 2018) [11]. The 2022 ACR/EULAR classification criteria for GPA, MPA and EGPA, which represent the validated version of the DCVAS criteria, were published after the study period; their implications for our cohort are considered in the Discussion [12–14]. Patients with another class of vasculitis were not included, and patients whose records were deemed unusable were excluded.

ANCA detection was carried out by indirect immunofluorescence on neutrophils fixed with ethanol, then with formalin. Anti-MPO or anti-PR3 antigen specificity was identified by enzyme-linked immunosorbent assay (ELISA) or by immunodot. The Birmingham Vasculitis Activity Score version 3 (BVAS v3) [15] and the revised Five-Factor Score (FFS) [16] were calculated at diagnosis and at each flare. The Vasculitis Damage Index (VDI) [17] was calculated at the last clinical examination. Proportions are reported with 95% confidence intervals (CI) calculated by the Wilson score method. Data were entered and analysed using SPSS version 21.0 (IBM SPSS Statistics).

## Results

Our study included nineteen patients with GPA and eleven patients with EGPA. Median age at diagnosis was 54 years [40–60] for GPA and 52 years [42–58] for EGPA. The male/female sex ratio was 0.46 for GPA and 0.57 for EGPA. Smoking was noted in 26% of GPA cases and 28% of EGPA cases.

Late-onset asthma, diagnosed after twenty years of age, was constantly recorded in EGPA. The median time between the onset of late asthma and diagnosis was 66 months. Major hypereosinophilia was present in all EGPA patients, with a median count of 4900/mm³ (range 1820–35376/mm³).

Lung involvement, apart from asthma, was registered in all cases of GPA and EGPA. A polymorphism of respiratory signs was observed in GPA, comprising nodules (74%), excavations (63%) and alveolar haemorrhage (37%). In EGPA, radiological infiltrates predominated (64%), including one case of chronic eosinophilic pneumonia (Figure 1). Lung biopsy, performed in 73% of EGPA patients, showed inflammatory and granulomatous infiltrates with vasculitis.

**Figure 1.**
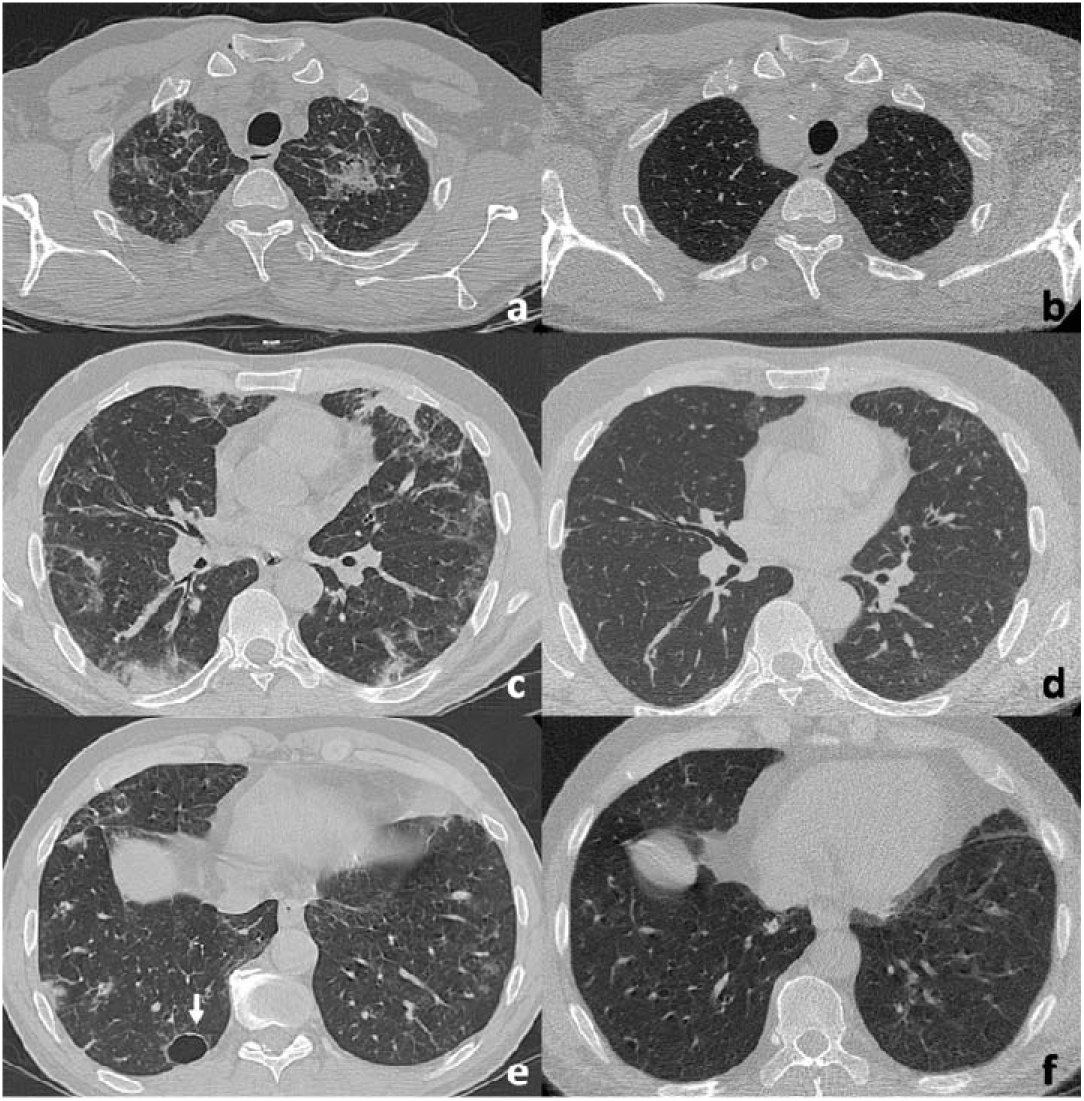
Eosinophilic pneumonia in a patient with EGPA.

Kidney involvement was noted in 53% of GPA and 27% of EGPA cases. In GPA, four patients had a pulmonary–renal syndrome and two had rapidly progressive glomerulonephritis complicated by advanced renal failure; kidney biopsy showed extracapillary or pauci-immune glomerulonephritis. In EGPA, patients had proteinuria and haematuria without renal failure, and kidney biopsy showed extracapillary glomerulonephritis in all cases.

Ear–nose–throat (ENT) involvement was noted in 79% of GPA cases, dominated by ulcerative crusty rhinitis (42%), sinusitis (37%) and conductive hearing loss (37%). Granuloma was identified in only 25% of nasal biopsies. Lesions were destructive in 32% of GPA cases, significantly associated with renal involvement (p=0.044). ENT involvement, noted in 91% of EGPA, comprised nasal polyposis in 36% of cases; an eosinophilic infiltrate was observed in 33% of EGPA nasal biopsies.

Peripheral neuropathy was noted in 64% of EGPA cases and was significantly correlated with ANCA positivity (p=0.013); it was also found in 25% of GPA cases. Electromyography revealed sensorimotor polyneuropathy more often than multiple mononeuropathy.

Cardiovascular involvement was frequent in EGPA (55% of cases, 95% CI 28–79), occurring only in women and often associated with negative ANCA (five of six patients). Heart involvement was noted in 26% of GPA cases, with prognostic value for conduction block.

Constitutional signs were registered in 68% of GPA and 82% of EGPA cases. Cutaneous, ophthalmic, digestive and thromboembolic disorders were less frequent in both diseases. At diagnosis, 47% of GPA and 55% of EGPA patients had at least one poor-prognosis factor (FFS ≥ 1).

Referring to the European Vasculitis Society (EUVAS) classification, we noted the absence of a localized form of GPA, one case of “early systemic” disease and two severe forms; the generalized form predominated (84% of GPA cases).

ANCA were constantly positive in GPA. By indirect immunofluorescence we identified perinuclear fluorescence (p-ANCA, 47%; Figure 2), cytoplasmic fluorescence (c-ANCA, 42%; Figure 3) and dual p-ANCA/c-ANCA fluorescence (5.5%); atypical x-ANCA were noted in 5.5%, checked on different samples. Antigenic specificity determined by ELISA or immunodot was anti-MPO (58%, 95% CI 36–77), anti-PR3 (26.5%, 95% CI 12–49) and dual MPO/PR3 specificity (11%, 95% CI 3–31) (Figure 4).

**Figure 2.**
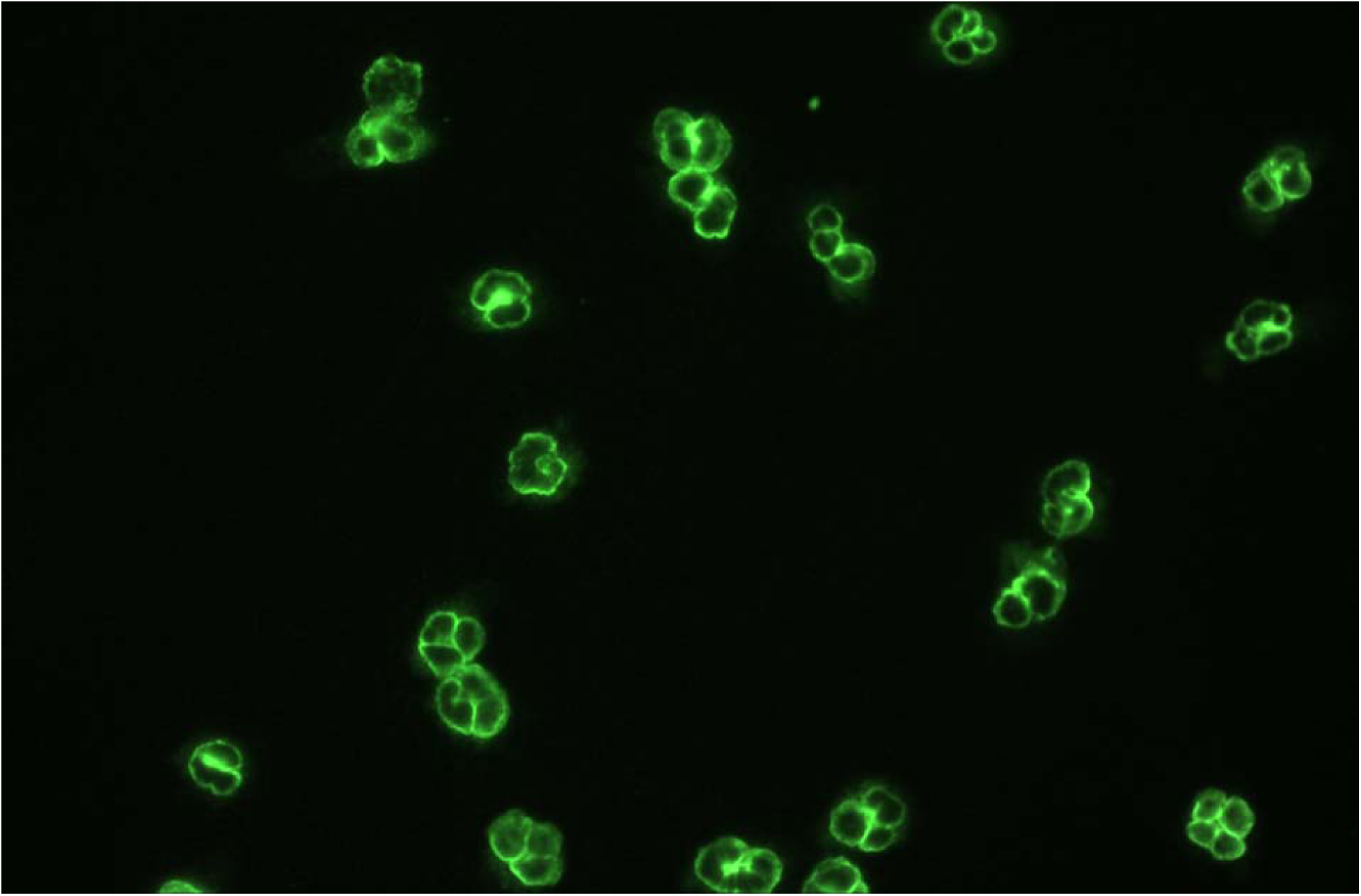
Perinuclear fluorescence (p-ANCA) by indirect immunofluorescence.

**Figure 3.**
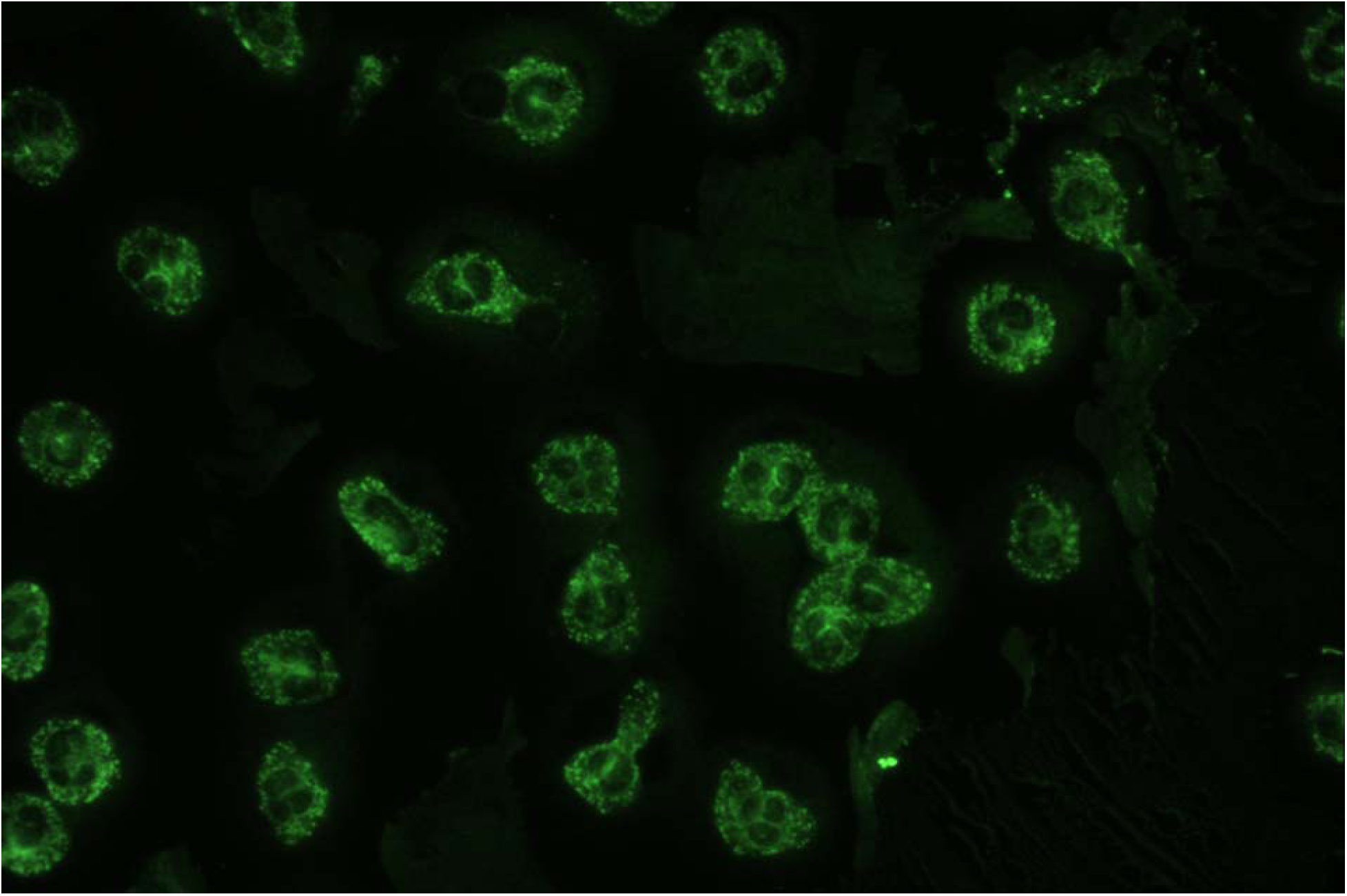
Cytoplasmic fluorescence (c-ANCA) by indirect immunofluorescence.

**Figure 4.**
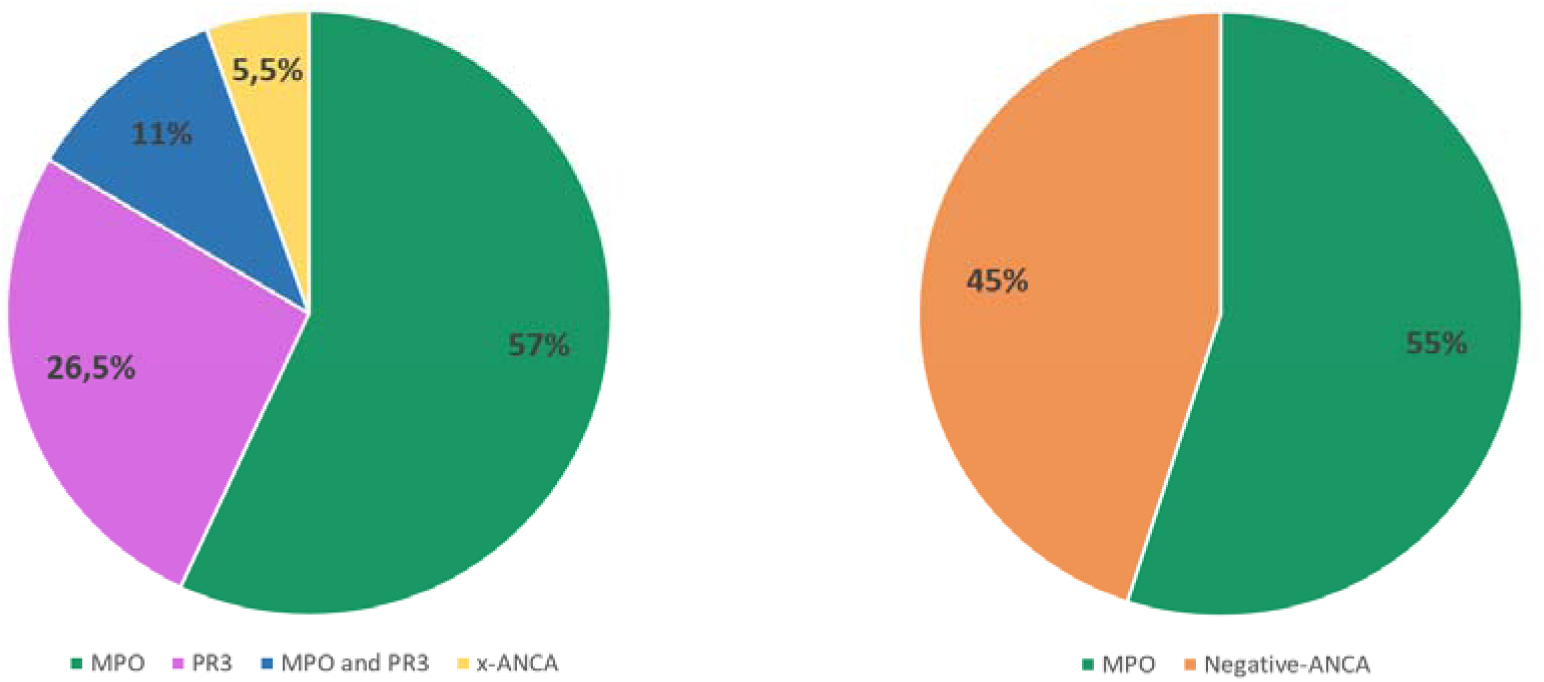
ANCA type and antigenic specificity in GPA (left) and EGPA (right).

ANCA were negative in 55% (95% CI 28–79) of EGPA cases. Immunofluorescence identified p-ANCA (18%) and c-ANCA fluorescence (27%); ANCA were directed against MPO in all positive cases (Figure 4). Testing for anti-glomerular basement membrane antibodies, performed in four patients with alveolar haemorrhage and one with pulmonary–renal syndrome, was negative in all cases.

Several clinical–immunological correlations were observed. In GPA, univariate analysis showed a different immunological profile by sex: anti-MPO antibodies were associated with female sex (p=0.005) and anti-PR3 with male sex (p=0.018). Anti-PR3 antibodies were also associated with smoking (p=0.019). In EGPA, peripheral neuropathy was associated with ANCA positivity (p=0.013); cardiac involvement was associated with negative ANCA in 83.3% of cases, and constitutional signs such as fever were more frequent when ANCA were negative (p=0.040). In the whole population, a non-significant trend toward an association between anti-MPO antibodies and alveolar haemorrhage was observed (p=0.053), and anti-PR3 antibodies were associated with a higher number of clinical features at diagnosis (p=0.006) and gastrointestinal involvement (p=0.033).

Therapeutic management followed the ACR/EULAR [18, 19] and/or the French Vasculitis Study Group recommendations [20]. As induction in GPA, all patients received high-dose oral prednisone (0.5–1 mg/kg/day), preceded by intravenous methylprednisolone pulses in 32% of cases; 84% received intravenous cyclophosphamide, and azathioprine, methotrexate and rituximab were each prescribed once. Plasma exchange was used in one case and haemodialysis in three. Maintenance consisted of azathioprine (53%), methotrexate (11%), rituximab (11%) or mycophenolate mofetil (5%). EGPA patients received intravenous methylprednisolone pulses (27%) and high-dose oral prednisone (100%), with intravenous cyclophosphamide (64%); maintenance was mainly azathioprine (64%).

After a median follow-up of 62 months, the relapse rate in EGPA was 36% (95% CI 15–65) with no deaths. In GPA, the relapse rate was 42% (95% CI 23–64) and mortality was 11% (95% CI 3–31) after a median follow-up of 74 months. Median BVAS v3 at GPA diagnosis was 13 [6–24], correlated with the number of flares (p=0.010), with a downward trend over successive flares (Figure 5). Median BVAS v3 at EGPA diagnosis was 16 [10–17], also with a downward trend. At last examination, 74% (95% CI 51–88) of GPA and 55% (95% CI 28–79) of EGPA patients had disease- or treatment-related damage, with respective median VDI of 3 [0–12] and 2 [0–3]. Higher mortality in GPA was correlated with a higher BVAS v3 at diagnosis (p=0.012), cardiac conduction disorder (p=0.039) and neurological damage (p=0.050).

**Figure 5.**
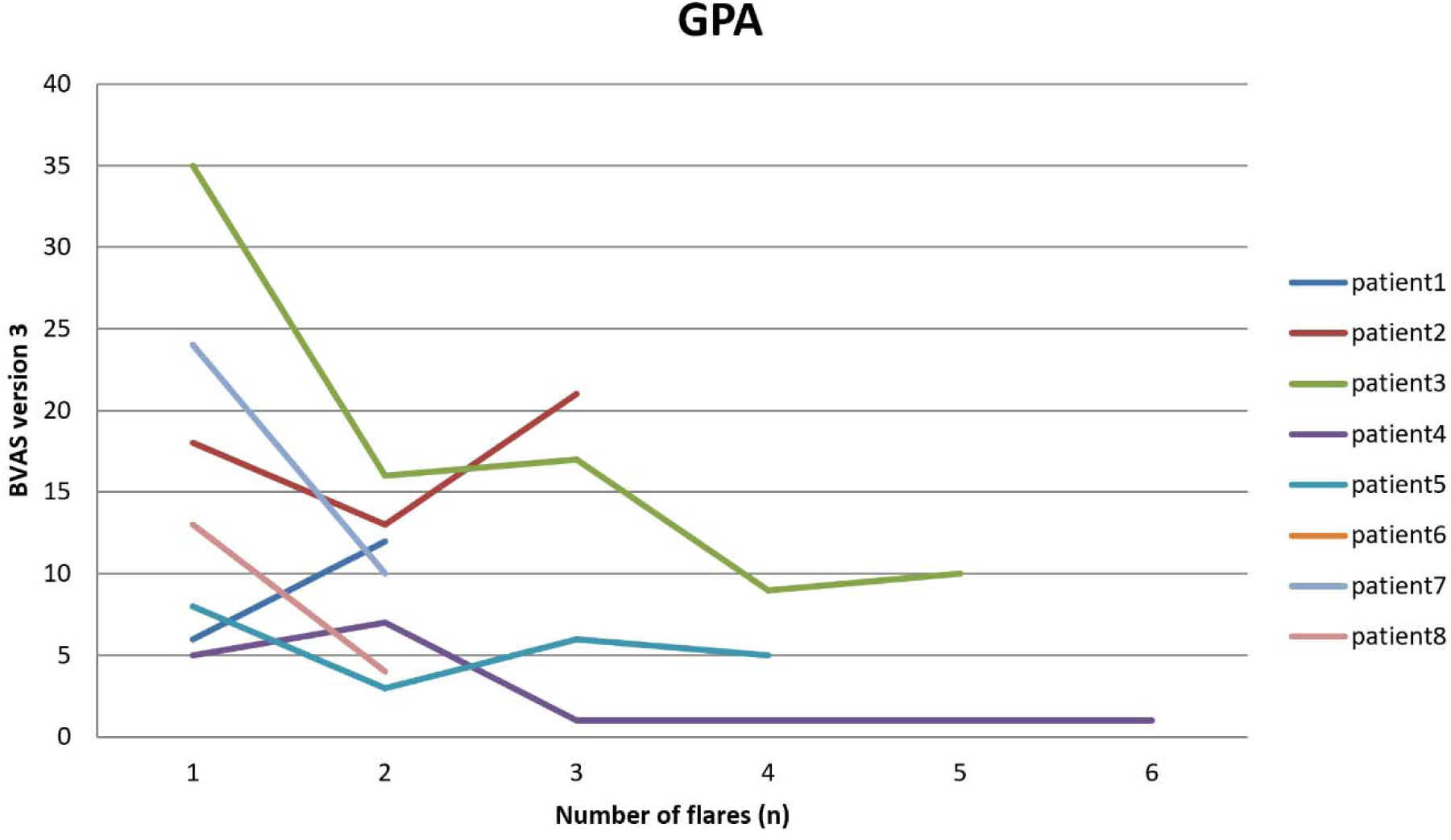
Downward trend in BVAS v3 over successive GPA flares.

## Discussion

The classification of AAV is shifting from a purely syndromic basis (GPA, MPA, EGPA) toward grouping by ANCA antigen specificity. A landmark genome-wide association study showed that the strongest genetic determinants of AAV track ANCA specificity rather than the clinical syndrome: PR3-ANCA disease with HLA-DP, SERPINA1 and PRTN3, and MPO-ANCA disease with This idea is now widely supported. Recent expert reviews debate whether AAV is better classified by clinical syndrome or by ANCA type, and they call for the 2022 ACR/EULAR criteria to be tested in under-represented populations, including in Africa [7, 8]. The 2022 criteria themselves incorporate ANCA specificity, assigning positive weight to PR3/c-ANCA and negative weight to MPO/p-ANCA for GPA [12–14]. Consistent with this, a recent multicentre analysis found good agreement between a specificity-based scheme and the 2022 criteria, while reclassifying some clinically defined GPA, particularly necrotizing non-granulomatous disease, as MPA [21]. Some experts have even proposed new categories, such as renal-limited AAV, which shows that how to classify these diseases is still debated [22–24].With its atypical serological distribution, our Tunisian cohort is a useful test case for this framework.

The median age at diagnosis (54 years for GPA, 52 for EGPA) was consistent with the literature [4, 5, 25], as was the disease expression, dominated by pulmonary, ear–nose–throat and constitutional involvement. We observed a female predominance in both diseases; most series report no clear sex predilection, although a slight male excess in GPA has been described [26, 27], and our finding may reflect the modest size and internal-medicine recruitment of the cohort. Renal involvement was less frequent than the approximately 75% reported in some GPA series [28, 29] and fell within the 16–22% range described for EGPA [5, 30]. Two local peculiarities deserve emphasis: the absence of localized GPA and the predominance of the generalized form [31].

The pathogenesis of AAV reflects gene–environment interaction and dysregulated innate and adaptive immunity [7, 32]. ANCA-mediated activation of primed neutrophils releases reactive oxygen species, lytic enzymes and neutrophil extracellular traps that injure the endothelium, with amplification through alternative-pathway complement (C5a) [7, 33]. The genetic architecture is specificity-dependent: the HLA-DP/SERPINA1/PRTN3 axis for PR3-AAV and HLA-DQ alleles for MPO-AAV, across European [6, 34] and East Asian [35] populations. Environmental exposures may shape specificity, with silica and some infections favouring MPO disease and nasal Staphylococcus aureus carriage favouring PR3 disease; in our cohort, smoking was associated with PR3-ANCA positivity (p=0.019) [36, 37]. This framework helps explain why ANCA specificity, and therefore disease expression, varies between populations.

In our GPA patients, ANCA were consistently positive, and MPO-ANCA outnumbered PR3-ANCA roughly two to one (58% vs 26.5%), the reverse of the PR3 predominance usually reported in GPA. ANCA type differs from one region to another: in the large international DCVAS study, most patients in Japan, China and southern Europe had MPO-ANCA, whereas most patients in northern Europe had PR3-ANCA [38], a pattern supported by genetic studies from East Asia [35]. Tunisia lies on the southern, Mediterranean side of this gradient, so a high proportion of MPO-ANCA in our patients is expected and biologically plausible, since MPO disease is linked to HLA-DQ genes that are common in these populations [6, 7].

This interpretation must be reconciled with a discrepancy: a Tunisian renal cohort from Charles Nicolle Hospital reported PR3 predominance (75.6%) among GPA patients [39], the opposite of our finding. The divergence between two Tunisian series is instructive rather than contradictory, most plausibly reflecting recruitment and specialty bias (a nephrology-based, biopsy-proven glomerulonephritis cohort versus our internal-medicine systemic cohort), together with differences in antigen-specific assay sensitivity (ELISA versus immunodot) and small sample sizes. A recent Eastern-Mediterranean cohort from Türkiye likewise reported a PR3-leaning distribution [40], underscoring genuine regional heterogeneity. These conflicting data, with ongoing ANCA characterisation at our own centre [41], argue that the true distribution of ANCA specificity in North Africa and the Mediterranean can only be resolved by larger, harmonised, multicentre studies.

Antigen specificity also shaped the disease pattern. MPO-ANCA disease more often affects the kidney and lung, whereas PR3-ANCA disease tends to involve more organs and to be more severe and relapse-prone [42, 43]. Our data fit this pattern, MPO-ANCA showed a trend toward an association with alveolar haemorrhage that did not reach significance (p=0.053), and a significant association with female sex (p=0.005). PR3-ANCA was associated with male sex (p=0.018), a higher number of clinical features (p=0.006) and gastrointestinal involvement (p=0.033). PR3-ANCA disease also tends to relapse more often: a meta-analysis found that PR3-ANCA independently predicts relapse (hazard ratio 1.69) [44], and other studies show that persistent PR3-ANCA and the return of B cells often precede relapse after rituximab [45]. The multi-organ PR3 pattern we observed therefore warrants closer monitoring, though our numbers are small. Dual MPO/PR3 positivity occurred in two GPA patients (11%), detected by immunodot in one and confirmed by ELISA in the other [46]. Double positivity is often linked to drug-induced disease (propylthiouracil, cocaine/levamisole) or to overlap with lupus or anti-GBM disease, and its prognostic value is uncertain [47]; such cases therefore warrant a search for a secondary cause and confirmation with an antigen-specific assay.

EGPA was ANCA-negative in 55% of patients and, when positive, exclusively anti-MPO. EGPA is now understood as two partially distinct entities: a genome-wide association study showed that MPO-ANCA-positive EGPA shares an HLA-DQ signal with MPO-AAV and behaves as a small-vessel vasculitis, whereas ANCA-negative EGPA carries a mucosal/barrier genetic signature and behaves as an eosinophilic, interleukin-5-driven tissue disease [48]. This division, formalised in the first EGPA-specific evidence-based guideline [49], matches the clinical features: vasculitic (glomerular, cutaneous and peripheral-nerve) manifestations in ANCA-positive disease, and cardiac and pulmonary-infiltrative disease in ANCA-negative disease [27, 50, 51]. Our findings fit this model: peripheral neuropathy was associated with ANCA positivity (p=0.013) and cardiac involvement clustered in ANCA-negative patients (five of six).

After a median follow-up of 62 (EGPA) and 74 (GPA) months, relapse rates were 36% and 42%, within the 30–50% reported internationally [44]. GPA mortality was 11% and was associated with a higher baseline BVAS v3 (p=0.012), conduction block (p=0.039) and neurological damage (p=0.050); the prognostic weight of cardiopulmonary disease is echoed by a regional cohort in which all deaths occurred in patients with pulmonary involvement [40]. Frequent damage (VDI ≥ 1 in 74% of GPA and 55% of EGPA) reflects both disease activity and the toxicity of conventional immunosuppression. Management has since evolved beyond the glucocorticoid–cyclophosphamide era of our cohort: the 2022 EULAR update favours rituximab and reduced-dose glucocorticoids and incorporates the C5a-receptor inhibitor avacopan [52, 53], while interleukin-5-targeted biologics (mepolizumab, and benralizumab shown non-inferior to it in the MANDARA trial) have transformed EGPA care [49, 54, 55].

## Limitations

This study has important limitations. Its retrospective, single-centre design and small sample size limit statistical power; all analyses are therefore exploratory and hypothesis-generating, no correction for multiple comparisons was applied, and the wide confidence intervals around our proportions reflect this uncertainty. Diagnosis spanned three classification systems, and ANCA were tested by both ELISA and immunodot; as antigen specificity was the principal variable, this assay heterogeneity is a particular limitation.

## Conclusion

This study provides a detailed clinical and immunological profile of GPA and EGPA in a Tunisian cohort and contributes data from a region where AAV remains under-reported. Two observations stand out: a predominance of MPO-ANCA among patients with GPA, at variance with the PR3 predominance classically described and with another Tunisian renal cohort, and a notable frequency of dual PR3/MPO positivity. These findings are compatible with a North–South gradient in ANCA specificity along the Mediterranean rim, but they must also be read in light of evolving classification: under the 2022 ACR/EULAR criteria, MPO-positive, necrotizing-predominant disease may be reclassified toward MPA, and the optimal grouping of AAV by ANCA specificity (PR3-versus MPO-AAV, with EGPA stratified by ANCA status) remains an open and biologically grounded question [6–8, 21]. Rather than resolving these issues, our cohort defines them locally and argues for larger, harmonised, multicentre Tunisian, Mediterranean and international studies integrating immunological, genetic and environmental data, and for formal validation of the 2022 criteria in African populations.

## What is already known on this topic

GPA and EGPA are among the rarest systemic vasculitides, characterised by necrotizing and granulomatous vasculitis and the inconstant positivity of ANCA, with wide variability in their clinical and immunological profile. Few North African studies have characterised their features in this region.

## What this study adds

In this Tunisian cohort, MPO-ANCA predominated even in GPA and dual PR3/MPO positivity was relatively frequent, supporting a probable North–South gradient in ANCA specificity while highlighting how the 2022 ACR/EULAR criteria may reclassify such cases. The findings strengthen the rationale for an ANCA-specificity-based approach to AAV classification and prognostication and for larger multicentre studies in the region [6, 8, 21].

## Data Availability

All data produced in the present study are available upon reasonable request to the authors

## Ethics approval

This study was conducted in accordance with the principles of the Declaration of Helsinki and was approved by the Institutional Ethics Committee of Mongi Slim Hospital, Tunis, Tunisia. Given the retrospective nature of the study and the use of anonymized data, the requirement for individual informed consent was waived. No specific ethics approval/reference number was assigned by the committee.

## Authors’ Confirmation

All authors confirm that they have contributed substantially to the conception and design of the study, acquisition, analysis, and interpretation of the data, and/or drafting and critical revision of the manuscript.

## Declaration of AI use

During the preparation of this manuscript, the authors used ChatGPT (OpenAI) solely for language editing, grammar correction, and improvement of English expression.

